# The variability of nociplastic and central pain definition in literature: a scoping review

**DOI:** 10.1101/2022.09.23.22279813

**Authors:** Michele Margelli, Bonas Enrico, Obinu Giovanni, De Marco Gianluca, Baroni Andrea, Sartorio Carlo

**Affiliations:** Faculty of Medicine and Surgery, Department of Clinical Science and Translation Medicine, University of Rome Tor Vergata, Roma, Italy; School of Physiotherapy, University of Ferrara, Ferrara, Italy; Studio Andreotti-Margelli Terapika, Ferrara, Italy; School of Physiotherapy, University of Ferrara; Department of Biomedical and Neuromotor Sciences (DIBINEM), University of Bologna, Bologna, Italy; Department of Neuroscience and Rehabilitation, Ferrara University Hospital, Via Aldo Moro 8, Ferrara, Cona, Italy

## Abstract

**Background:** Currently, the concept of pain is widely discussed in the scientific community, particularly chronic musculoskeletal pain.

One important classification concerns the mechanisms of pain generation, according to which pain is divided into nociceptive pain, neuropathic pain, nociplastic pain (NP), and central sensitization (CS).

Often the terms nociplastic pain and central sensitization are used incorrectly, as synonyms, or improperly; this can make data transmission complicated.

The aim of this review will be to provide a clearer overview of the concept of pain in the scientific literature, describe the variability on the use of the terms nociplastic pain and central sensitization, and describe the mechanisms in relation to musculoskeletal pain syndromes.

**Inclusion Criteria:** Every study describing the mechanisms of nociplastic pain or central sensitization in a population of adults (> 18 years old), with acute or chronic musculoskeletal pain, in one or more anatomic regions. This scoping review will consider studies conducted in any context. Articles in English or Italian will be considered.

**Methods:** The proposed scoping review will be conducted in accordance with the Joanna Briggs Institute methodology (JBI) for scoping reviews.

The search will be carried out on 5 databases: MEDLINE, Cochrane Central, Scopus, Embase, and PEDro.

Selection and data extraction will be conducted by two blind independent researchers and inconsistencies will be resolved by a third reviewer.

The results will be presented in a schematic, tabular and descriptive format that will line up with the objectives and scope of the review.

**Conclusions:** This will be the first scoping review to provide a comprehensive overview of the topic. The results will add meaningful information for clinicians. Furthermore, any knowledge gaps of the topic will be identified. The results of this research will be published in a peer-reviewed journal and will be presented at relevant (inter)national scientific events.

## BACKGROUND

Nowadays the concept of pain is widely discussed in the scientific community, especially musculoskeletal chronic pain, indicated as the worldwide widespread pathology that leads to disability and enormous socio-economic burden (1).

In particular musculoskeletal chronic pain comprises a major public health problem in developed countries owing to high prevalence rates and considerable burden in terms of medical costs, work disability, and reduced quality of life (2), but actually data of the prevalence of most musculoskeletal disease are poor (3).

The relative estimate of the prevalence of its population varies widely based on the definition of almost (4), more than about 20% of the European adult population suffering from chronic pain (5), in particular musculoskeletal pain affects between 13.5% and 47% of the general population, with CWP (Chronic widespread Pain) prevalence varying between 11.4% and 24% (6).

For this reason, it is considered a major priority in the field of research by the public health institutions such as the WHO (World Health Organization) in terms of impact on economic growth, socio-sanitary and its primary and specialist assistance services (7).

Based on the most recent definitions of pain proposed by the International Association for the Study of Pain (IASP), pain is defined as “An unpleasant sensory and emotional experience associated with, or resembling that associated with, actual or potential tissue damage” (8).

Today the new ICD-11 classification introduces the concept of chronic primary and secondary musculoskeletal pain, and integrates the biomedical axis with the psychological and social axes that comprise the complex experience of chronic musculoskeletal pain.(9)

Another classification is about the mechanisms of pain generation, or “pain generators’’ (10) according to which pain is divided into nociceptive, neuropathic pain(11), nociplastic pain (NP), and Central sensitization (CS), this term is often associated as chronic pain (12).

A combination of one or more types of pain may occur. (11).

Unfortunately, today the multitude of terms, often used improperly, can confuse or make data transmission complicated (7), so this scoping review will aim to provide a clearer overview of the concept of pain in the scientific literature, describe variability about nociplastic and CS terms use, map the available literature about these terms and describing the mechanisms of central pain (nociplastic and central sensitization) in relation to musculoskeletal pain syndromes.

## Review question

Are the terms nociplastic and central sensitization used synonymously or do they mean distinct realities?

The main objectives of this study will be:

1. To map the available literature regarding the use of terms describing the mechanisms of central-type pain (nociplastic, central sensitization), in relation to musculoskeletal pain syndromes.
2. Describe variability in the use of the term nociplastic.
3. Describe variability in the use of the term central sensitization.
4. Check the semantic overlap of the terms nociplastic and central sensitization, and in case of discordance propose a shareable terminological usage for each term.

## Methods

### Study design and protocol

The proposed scoping review will be conducted in accordance with the Joanna Briggs Institute methodology (JBI) for scoping reviews.

The Preferred Reporting Items for Systematic reviews and Meta-Analyses extension for Scoping Reviews (PRISMA-ScR) Checklist for reporting will be used, and it is priori registered at Medxriv (https://www.medrxiv.org).

### Search strategy

The search will be carried out on 6 databases: MEDLINE, Cochrane Central, Scopus, Embase, and PEDro. Studies will also be included searching in the bibliography of relevant revisions, and in Google Scholar. Further research of Gray literature will be carried out through open gray.eu, a multidisciplinary European database. Keywords inserted are: nociplastic, central sensitization, pain, musculoskeletal.

As recommended in all JBI types of reviews and PRISMA-S, a three-step search strategy is to be utilized.

1. The first step is an initial limited search of an appropriate online databases relevant to the topic (MEDLINE). This initial search is then followed by an analysis of the text words contained in the title and abstract of retrieved papers, and of the index terms used to describe the articles.
2. The second step concerns a search using all identified keywords and index terms should then be undertaken across all included databases.
3. The third step, the reference list of identified reports and articles should be searched for additional sources.

The search strategies were peer-reviewed by an experienced librarian and were further refined through team discussion. No search limitations and filters applied (language and time). Reviewers’ intent to contact authors of primary sources or reviews for further information. Complete search strategy for Medline is included as an appendix 1 to the protocol. Search strategy will be adapted to be used in other databases.

### Inclusion criteria

We will follow the acronym PCC to describe elements of the inclusion criteria:

#### Population

This scoping review will consider studies with adults (> 18 years old) with musculoskeletal acute and chronic pain, in 1 or more anatomic regions

#### Concept

This scoping review will consider research studies that describe nociplastic or central sensitization mechanisms.

#### Context

This scoping review will consider studies conducted in any context.

#### Sources

This scoping review will consider any study designs or publication type for inclusion. No date and geographical limits will be used. We will consider articles in English and Italian. Studies that do not meet the above-stated Population-Concept-Context (PCC) criteria or which provide insufficient information will be excluded. Any work that does not contain a definition or reference of the term nociplastic or central sensitization will also be excluded.

### Study selection

For the selection process, the first thing to do is to select a sample of 25 title/abstracts that will be analyzed from the entire team using eligibility criteria and definitions/elaboration document. Then, the team will discuss discrepancies and make modifications to the eligibility criteria and definitions/elaboration document. The team will only start screening when 75% (or greater) of agreement between reviewers will be achieved. To start the screening, we will use the Rayyan online software (https://www.rayyan.ai).

The selection phase will begin assessing all the titles and abstracts of the studies retrieved using the search strategy and those from additional sources. These studies will be screened independently by two review authors, to identify those that potentially meet the inclusion criteria. Any disagreement will be solved by two reviewer consensus. Subsequently, the full text of these potentially eligible studies will be independently assessed for eligibility by the two reviewers. The reasons for excluding articles will be recorded. We developed a google form (charting table) containing the elements to standardize the selection.

There should be a narrative description of the selection process accompanied by a flowchart of review process (from the PRISMA-ScR statement).

### Data extraction

Extraction module (in appendix B) will be reviewed, before the implementation, by the research team and pre-tested to ensure that the form accurately captures the information. Modifications will be detailed in the full scoping review.

Data extraction will be conducted by two blind independent researchers and inconsistencies will be resolved by a third reviewer.

### Data items

Key information will be described in a charting table with the description of: Author; Publication year; Place of study’s conduct; Setting of study’s conducts; Methodology/ type of study; Aims of the study; Population, Characteristics from which patients are extracted, including gender and age, social variables; Concept: Definition of nociplastic pain or central sensitization. Tools used to assess the possible presence of nociplastic pain or central sensitization mechanisms.

### Critical appraisal of individual sources of evidence

Not provided.

### Data management

As a scoping review, the purpose is to aggregate the findings and to present an overview of the research rather than to evaluate the quality of the individual studies. The results will be presented as a map of data, which are extracted from different documents. These will be included in a schematic, tabular and descriptive format that will line up with the objectives and scope of the review. Descriptive analysis will consist a distribution of the evidence sources by year or period of publication, countries of origin, area of intervention (clinical, political, educational, etc.) and research methods. Results will be presented such as: population (gender, age), nociplastic pain, central sensitization and the tools used to assess them. An overall classification of nociplastic pain and central sensitization with narrative description and table will be proposed.

## Data Availability

All data produced in the present study are available upon reasonable request to the authors

## Data Availability

Due to the nature of this research, participants of this study did not agree for their data to be shared publicly, so supporting data is not available.

## Appendix A Complete search strategy

(“nociplastic”[All Fields] OR ((“central”[All Fields] OR “centrally”[All Fields] OR “centrals”[All Fields]) AND (“sensitisation”[All Fields] OR “sensitisations”[All Fields] OR “sensitise”[All Fields] OR “sensitised”[All Fields] OR “sensitiser”[All Fields] OR “sensitisers”[All Fields] OR “sensitises”[All Fields] OR “sensitising”[All Fields] OR “sensitization”[All Fields] OR “sensitizations”[All Fields] OR “sensitize”[All Fields] OR “sensitized”[All Fields] OR “sensitizer”[All Fields] OR “sensitizers”[All Fields] OR “sensitizes”[All Fields] OR “sensitizing”[All Fields])) OR “central pain”[All Fields]) AND (“muscoloskeletal”[All Fields] OR (“musculoskeletal system”[MeSH Terms] OR (“musculoskeletal”[All Fields] AND “system”[All Fields]) OR “musculoskeletal system”[All Fields] OR “musculoskeletal”[All Fields]) OR “MSK”[All Fields]) AND (“pain”[MeSH Terms] OR “pain”[All Fields])

## Reference

1. James SL, Abate D, Abate KH, Abay SM, Abbafati C, Abbasi N, et al. Global, regional, and national incidence, prevalence, and years lived with disability for 354 diseases and injuries for 195 countries and territories, 1990–2017: a systematic analysis for the Global Burden of Disease Study 2017. The Lancet. novembre 2018;392(10159):1789–858.

2. Wijnhoven HAH, de Vet HCW, Picavet HSJ. Prevalence of Musculoskeletal Disorders Is Systematically Higher in Women Than in Men. Clin J Pain. ottobre 2006;22(8):717–24.

3. Picavet HSJ. Prevalence of self reported musculoskeletal diseases is high. Ann Rheum Dis. luglio 2003;62(7):644–50.

4. Mills SEE, Nicolson KP, Smith BH. Chronic pain: a review of its epidemiology and associated factors in population-based studies. Br J Anaesth. agosto 2019;123(2):e273–83.

5. van Hecke O, Torrance N, Smith BH. Chronic pain epidemiology and its clinical relevance. Br J Anaesth. luglio 2013;111(1):13–8.

6. Cimmino MA, Ferrone C, Cutolo M. Epidemiology of chronic musculoskeletal pain. Best Pract Res Clin Rheumatol. aprile 2011;25(2):173–83.

7. Nijs J, Lahousse A, Kapreli E, Bilika P, Saraçoğlu İ, Malfliet A, et al. Nociplastic Pain Criteria or Recognition of Central Sensitization? Pain Phenotyping in the Past, Present and Future. J Clin Med. gennaio 2021;10(15):3203.

8. Raja SN, Carr DB, Cohen M, Finnerup NB, Flor H, Gibson S, et al. The revised International Association for the Study of Pain definition of pain: concepts, challenges, and compromises. Pain. settembre 2020;161(9):1976–82.

9. Perrot S, Cohen M, Barke A, Korwisi B, Rief W, Treede RD, et al. The IASP classification of chronic pain for ICD-11: chronic secondary musculoskeletal pain. Pain. gennaio 2019;160(1):77–82.

10. Bonezzi C, Fornasari D, Cricelli C, Magni A, Ventriglia G. Not All Pain is Created Equal: Basic Definitions and Diagnostic Work-Up. Pain Ther. dicembre 2020;9(S1):1–15.

11. Trouvin AP, Perrot S. New concepts of pain. Best Pract Res Clin Rheumatol. giugno 2019;33(3):101415.

12. Nijs J, Torres-Cueco R, van Wilgen CP, Girbes EL, Struyf F, Roussel N, et al. Applying modern pain neuroscience in clinical practice: criteria for the classification of central sensitization pain. Pain Physician. ottobre 2014;17(5):447–57.

