## Appendix B for "The variability of nociplastic and central pain definition in literature: a scoping review"

|  | Elements |
| --- | --- |
| Article Details  -Article title  -Author(s)  -Year of pubblication  -Origin/country of origin  -Setting context  -Methodology/Methods  -Languages |  |
| Population  -Numbers  -Mean age  -Class of age  -Male  -Female  -Pathology (cronic and acute pain)  -Variables Factors |  |
| Concept  -Definition of nociplastic pain or central sensitization  -Tools used to assess the possibile presence of nociplastic pain or central sensitization mechanism |  |
